# Time to First-Line Antiretroviral Therapy Failure and Its Predictors among People Living with HIV in Tanzania

**DOI:** 10.64898/2026.03.13.26348346

**Authors:** Raphael Z. Sangeda, Henry Gaspar Bahati, Natorias Mgaya Salvatory, James Mwakyomo, Veryeh Sambu, Prosper Njau

**Affiliations:** Department of Pharmaceutical Microbiology, Muhimbili University of Health and Allied Sciences, Dar es Salaam, Tanzania; Department of Natural Product Development and Formulation, Muhimbili University of Health and Allied Sciences, Dar es Salaam, Tanzania; National AIDS, Sexually Transmitted Infections and Hepatitis Control Programme, Dodoma, Tanzania

**Keywords:** HIV, antiretroviral therapy, first-line treatment failure, time-to-event analysis, Cox proportional hazards, dolutegravir, Tanzania

## Abstract

**Introduction:** Sustaining long-term viral suppression among people living with HIV (PLHIV) remains a major public health challenge in sub-Saharan Africa, despite widespread access to antiretroviral therapy (ART). Evidence on time to first-line ART failure and its predictors at a national scale remains limited, particularly for dolutegravir (DTG)-based regimens. We aimed to estimate the time to first-line ART failure and identify associated predictors among PLHIV in Tanzania using national programmatic data.

**Methods:** We conducted a retrospective cohort study using routinely collected data from the National Care and Treatment Clinic database (CTC-2) in Tanzania. The analysis included PLHIV aged ≥11 years who initiated first-line ART between January 2017 and December 2021 and had at least six months of follow-up. Time to first-line ART failure was defined as the duration from ART initiation to the first documented virological failure (viral load ≥1,000 copies/mL). Kaplan-Meier methods were used to estimate failure-free survival, and Cox proportional hazards models were used to identify predictors of failure. Non-proportional hazards for DTG-based regimens were addressed using an extended Cox model with a time-varying coefficient.

**Results:** The final analytic cohort comprised 36,764 individuals and 789 first-line treatment failure events. Median follow-up time varied across regimen groups. Failure-free survival differed significantly by regimen anchor (log-rank p<0.001). In multivariable Cox models, age and gender were significantly associated with treatment failure. DTG-based regimens demonstrated a time-varying effect: compared with non-DTG regimens, DTG was associated with a substantially lower hazard of failure early after initiation, with the protective effect attenuating over time. Estimated hazard ratios for DTG versus non-DTG regimens were 0.37, 0.67, and 1.22 at 6, 12, and 24 months of follow-up, respectively.

**Conclusions:** In this large national cohort, the risk of first-line ART failure varied by regimen and patient characteristics. DTG-based regimens showed strong early protection against failure, but this effect diminished over time, highlighting the importance of continued virological monitoring after ART initiation. Time-to-event analyses using routine programmatic data provide important evidence for optimizing ART delivery and informing HIV programme decisions in Tanzania and similar settings.

## 1.0 Introduction

Human immunodeficiency virus (HIV) remains a major global public health challenge, particularly in sub-Saharan Africa. In 2024, an estimated 40.8 million people were living with HIV worldwide, an increase from 40.4 million in 2023, mainly reflecting a continued decline in HIV-related mortality. The annual HIV-related deaths decreased from approximately 650,000 in 2023 to 630,000 in 2024. Sub-Saharan Africa bears the greatest burden, accounting for more than half of all people living with HIV (PLHIV), with an estimated 26.3 million affected individuals [1].

In Tanzania, HIV continues to impose a substantial public health burden, despite major progress in expanding access to antiretroviral therapy (ART). The 2022-2023 Tanzania HIV Impact Survey reported an adult HIV prevalence of 4.4% [2]. Widespread ART scale-up has resulted in marked reductions in morbidity and mortality; however, treatment failure remains a persistent challenge in achieving durable viral suppression and sustained epidemic control.

According to the 2019 national HIV treatment guidelines, the recommended first-line ART regimens for adults and adolescents consist of two nucleoside reverse transcriptase inhibitors (NRTIs), including tenofovir (TDF), lamivudine (3TC), zidovudine (AZT), or abacavir (ABC), combined with either a non-nucleoside reverse transcriptase inhibitor (NNRTI), most commonly efavirenz (EFV), or an integrase strand transfer inhibitor (INSTI), primarily dolutegravir (DTG). For children, regimen selection varies by age, with boosted protease inhibitors (PIs) forming the backbone of the second-line therapy. In contrast, third-line regimens include darunavir/ritonavir or advanced INSTIs such as raltegravir [3,4].

Despite the availability of effective ART regimens, first-line treatment failure is a significant concern. A cohort study of Tanzanian adults receiving NNRTI-based therapy reported a virological failure rate of 14.9% within 24 weeks of treatment initiation [5]. Similarly, immunological failure was observed in 17.1% of patients followed for a median of 29 months at Bugando Hospital in Mwanza [6]. High levels of resistance-associated mutations, particularly affecting NNRTIs and NRTIs, have been documented in patients with suspected treatment failure [7,8]. Among individuals receiving second-line ART, the virological failure rates approach 30%, with an incidence of approximately 92.8 per 1,000 person-years [9].

Treatment failure, whether virological, immunological, or clinical, has profound implications for patient care and public health. It accelerates disease progression, increases the risk of opportunistic infections, elevates healthcare costs, and contributes to onward HIV transmission. Virological failure, defined as a persistent viral load ≥1,000 copies/mL despite ART, is considered the most reliable indicator of treatment failure. Identified risk factors include poor adherence, late ART initiation, high baseline viral load, low CD4 cell count, drug resistance, and tuberculosis co-infection [5–7,9–14].

Understanding the timing of first-line ART failure and its predictors is essential for optimizing HIV care and informing timely treatment decision-making. Time-to-event analyses provide insights into the durability of first-line regimens, identify populations at a higher risk of failure, and guide decisions regarding regimen switching. Previous studies in Tanzania and neighboring countries have reported predictors of treatment failure, including gender, tuberculosis co-infection, advanced WHO clinical stage, and low baseline CD4 counts [9,11,12,14,15]. However, much of the available evidence is derived from pediatric cohorts, adolescents, or patients on second-line therapy, highlighting a gap in nationally representative analyses of first-line ART durability in adults.

The National AIDS, Sexually Transmitted Infections, and Hepatitis Control Programme (NASHCoP) maintains the national Care and Treatment Clinic (CTC-2) database, a comprehensive electronic system that captures longitudinal demographic, clinical, laboratory, and treatment data from HIV care and treatment facilities across Tanzania. Leveraging this database provides a unique opportunity to generate real-world, population-level evidence on ART outcomes in Tanzania.

Therefore, this study aimed to determine the time to first-line ART failure and its predictors among people living with HIV in Tanzania using the national CTC-2 database. The findings are intended to inform clinical practice, programmatic interventions, and policy decisions aimed at sustaining long-term treatment success and strengthening HIV control.

## 2.0 Methods

### 2.1 Study Design

This retrospective cohort study used routinely collected national programmatic data from the National AIDS, Sexually Transmitted Infections, and Hepatitis Control Programme (NASHCoP). The primary objective was to estimate the time to first-line antiretroviral therapy (ART) failure and identify predictors of failure among people living with HIV (PLHIV) in Tanzania. A time-to-event analytical framework was applied to capture both the duration of first-line ART and the evolving risk of treatment failure during follow-up.

### 2.2 Study Area

This study utilized data from the National CTC-2 database, which consolidates longitudinal clinical, demographic, laboratory, and treatment information from HIV care and treatment facilities across Tanzania. Tanzania has a generalized HIV epidemic, with an estimated 1.7 million PLHIV, approximately 1.4 million of whom receive ART. The national population exceeds 61 million, and the prevalence of HIV varies substantially across regions. The adult HIV prevalence was estimated to be 4.4% in 2023 [2].

### 2.3 Study Population

The study population comprised PLHIV aged 11 years and older who initiated first-line ART between 1 January 2017 and 31 December 2021 and were recorded in the CTC-2 database. Eligible participants were those who initiated nationally recommended first-line ART regimens with available baseline demographic and regimen information. Patients were excluded if essential baseline data, such as ART start date, age, or regimen details, were missing; if they were younger than 11 years at ART initiation; if they were transferred into care without baseline treatment records; or if they died, transferred out, or were lost to follow-up before completing at least six months of ART, thereby preventing reliable assessment of first-line treatment failure.

### 2.4 Sample Size and Sampling Procedure

Because of the size of the national CTC-2 database and computational requirements for time-to-event modelling, a simple random sample of approximately 50,000 individuals was drawn from the full registry before applying eligibility criteria. All patients meeting the eligibility criteria during the study period were identified in this dataset by applying predefined inclusion and exclusion criteria, yielding a final analytic cohort of 36,764 individuals.

### 2.5 Variables

The primary outcome was time to first-line ART failure, defined as the duration in days from ART initiation to the first documented virological failure. Virological failure was defined as a viral load of at least 1,000 copies/mL occurring at least 6 months after ART initiation, in accordance with the national and World Health Organization guidelines. Individuals who did not experience virological failure were censored on the date of their last recorded clinical contact.

The independent variables were selected a priori based on clinical relevance, data availability, and the existing literature. These included age at ART initiation (continuous), gender, and the region of residence. Treatment-related variables included the first-line ART regimen, categorized by the anchor drug as dolutegravir (DTG), efavirenz (EFV), nevirapine (NVP), or a protease inhibitor (PI), and the average pharmacy refill adherence expressed as a continuous percentage. The follow-up characteristics included the total duration of follow-up and the number of clinic visits.

Clinical staging variables, such as WHO stage, tuberculosis co-infection, opportunistic infections, baseline CD4 count, and baseline viral load, were not consistently available across the national dataset. Therefore, they were excluded from the multivariate analyses.

### 2.6 Data Collection and Management

Data were extracted from the CTC-2 database, which is populated using standardized CTC-1 clinical encounter forms completed at each clinic visit. Extracted variables included demographic characteristics, ART initiation dates, regimen histories, viral load test results, adherence measures derived from pharmacy refill data, and treatment outcomes.

Data management procedures included removing duplicate records, verifying temporal consistency, and excluding ineligible observations. Derived variables were created to facilitate the time-to-event analysis, including follow-up time, censoring indicators, regimen anchor classification, and failure events. All analyses were conducted using fully anonymized data sets.

### 2.7 Statistical Analysis

Baseline characteristics were summarized using medians with interquartile ranges for continuous variables and frequencies with percentages for categorical variables. Baseline distributions were described overall and by the first-line regimen anchor category.

Time-to-event analyses were conducted using Kaplan-Meier methods to estimate failure-free survival probabilities over time to first-line ART failure. Survival curves were compared across the regimen groups using the log-rank test.

Cox proportional hazards regression models were fitted to estimate crude hazard ratios (CHR) and adjusted hazard ratios (AHR) with 95% confidence intervals for predictors of ART failure. Variables with p-values below 0.25 in the univariable analyses were considered for inclusion in the multivariable models. The geographic region was included as a stratification variable to account for regional heterogeneity in the baseline hazards.

Evidence of non-proportional hazards for DTG-based regimens prompted the use of an extended Cox model incorporating a time-varying coefficient for DTG exposure that interacted with the log-transformed time. Hazard ratios (HRs) comparing DTG-based regimens with non-DTG regimens were estimated at 6, 12, and 24 months after ART initiation. Pharmacy refill adherence was calculated as the average proportion of days covered across the follow-up period. Because this measure incorporates information accumulated after treatment initiation, it was not included as a baseline predictor in the primary Cox regression models to avoid time-dependent bias. Adherence was therefore examined descriptively and only in sensitivity analyses.

The proportional hazards assumption was assessed using Schoenfeld residuals in the baseline Cox models. Formal proportional hazards testing was not applied to the models with time-varying coefficients because the assumption of proportional hazards did not hold. Statistical significance was defined as a two-sided p-value of < 0.05. All analyses were performed in R.

### 2.8 Ethical Considerations

Ethical approval was obtained from the Directorate of Research, Publications, and Innovation at Muhimbili University of Health and Allied Sciences (MUHAS) (Reference: DA.25/111/28/01/2021). Permission to access and analyze programmatic data was granted by NASHCoP. All data were anonymized prior to analysis, and no personal identifiers were retained in the dataset.

## 3.0 Results

### Study population and baseline characteristics

A total of 36,764 individuals initiating first-line antiretroviral therapy were included in the main analytical cohort, contributing to 789 first-line ART failure events during follow-up. Most participants initiated efavirenz (EFV)-based regimens, followed by dolutegravir (DTG), nevirapine (NVP), and protease inhibitor (PI)-based regimens (Table 1).

**Table 1:**
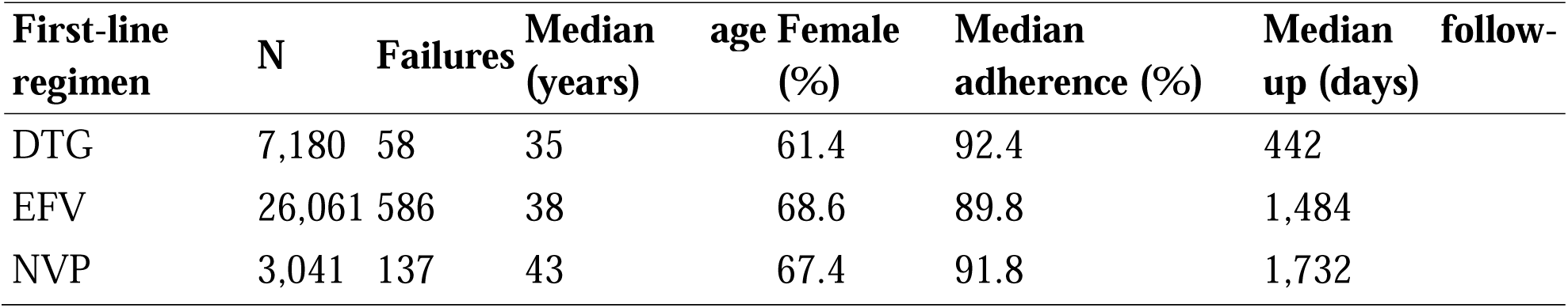
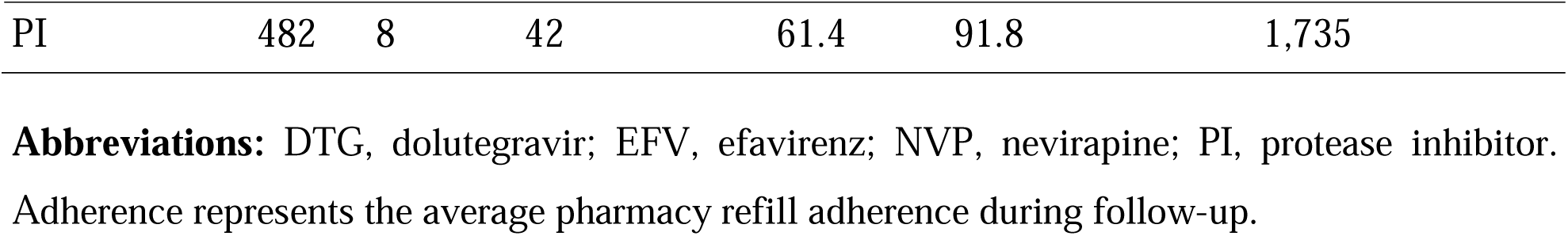
Baseline characteristics of the study population by first-line ART regimen.

The median age at ART initiation ranged from 35 years among DTG recipients to 43 years among those initiated on NVP-based regimens. Females accounted for 61.4% to 68.6% of patients across the regimen groups. The median adherence was high across all groups, exceeding 89%. In contrast, the median follow-up time varied substantially by regimen, with the shortest time for DTG (442 days) and the longest for PI and NVP regimens (approximately 1,700 days).

### Failure-free survival by first-line regimen

Failure-free survival over time by regimen anchor is shown in **Figure 1**.

**Figure 1:**
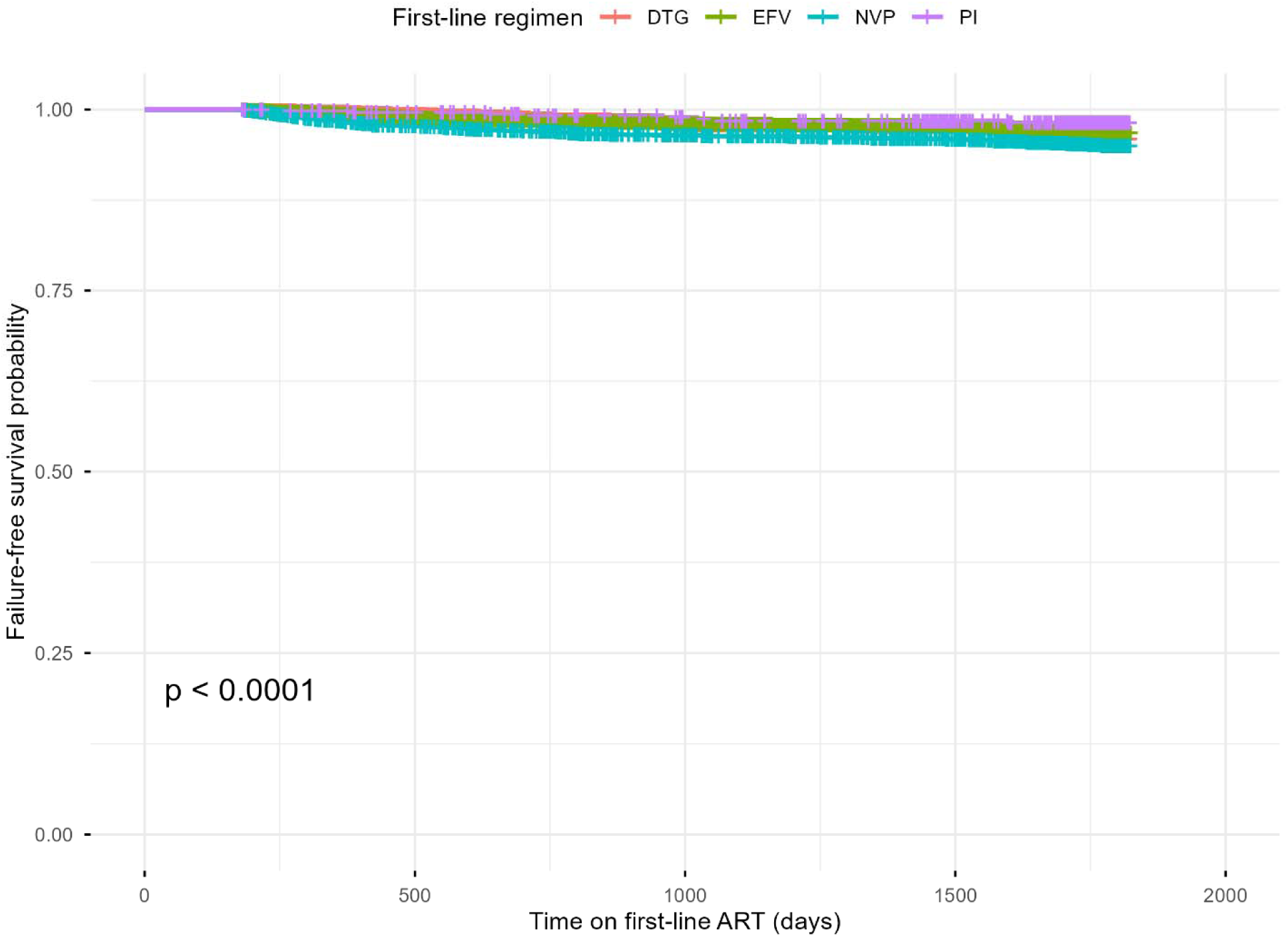
**Kaplan-Meier curves of failure-free survival by first-line ART regimens.**

The survival curves for DTG, EFV, NVP, and PI-based regimens largely overlapped throughout the follow-up period, with only modest absolute separation. Despite this visual overlap, the overall comparison across regimens was statistically significant (log-rank p < 0.0001), reflecting the large sample size and extended observation period rather than the marked clinical divergence between regimens.

### Cox proportional hazards models assuming baseline effects

The crude and adjusted Cox proportional hazards models for time to first-line ART failure are presented in Table 2.

**Table 2:**
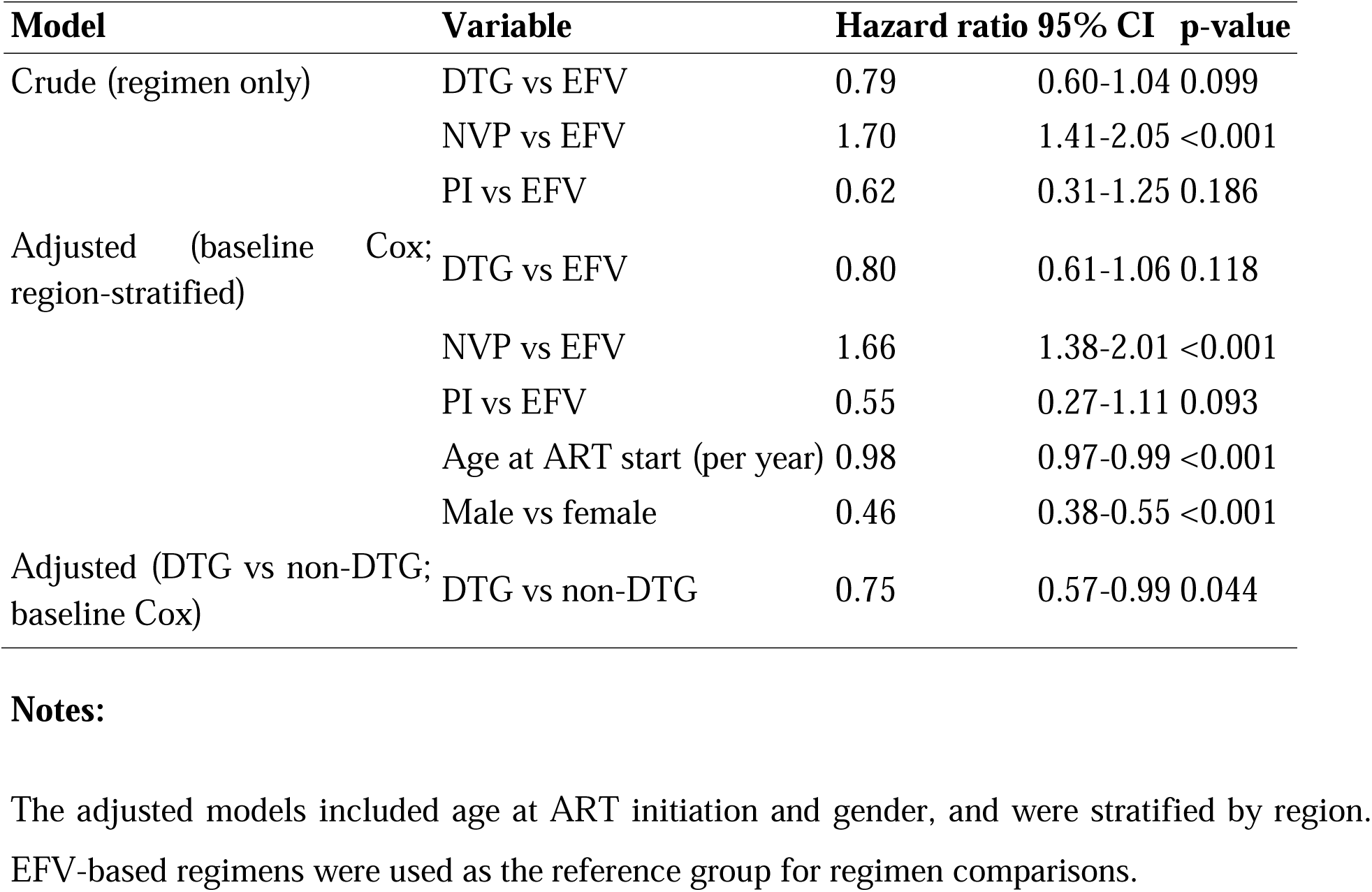
Crude and adjusted Cox proportional hazards models for time to first-line ART failure.

In crude analyses, DTG-based regimens were associated with a lower risk of failure than EFV-based regimens, although this association did not reach statistical significance. NVP-based regimens were associated with a significantly higher hazard of failure, whereas PI-based regimens showed no clear association, likely due to limited event counts.

In the adjusted models, controlling for age at ART initiation and gender and stratified by region, DTG-based regimens remained associated with a lower hazard of failure compared with non-DTG regimens, although the effect size was attenuated. Male gender was associated with a lower hazard of failure, whereas increasing age was associated with a modest reduction in hazard.

### Time-varying effect of DTG-based regimens

Assessment of the proportional hazards assumptions indicated a violation for DTG exposure, motivating the use of a Cox model with a time-varying DTG effect. Model fit statistics favored the time-varying specification compared to the baseline DTG model.

Time-specific hazard ratios derived from the time-varying model at 6, 12, and 24 months are shown in Table 3. In contrast, the continuous trajectory of the hazard ratio over time is illustrated in Figure 2.

**Figure 2:**
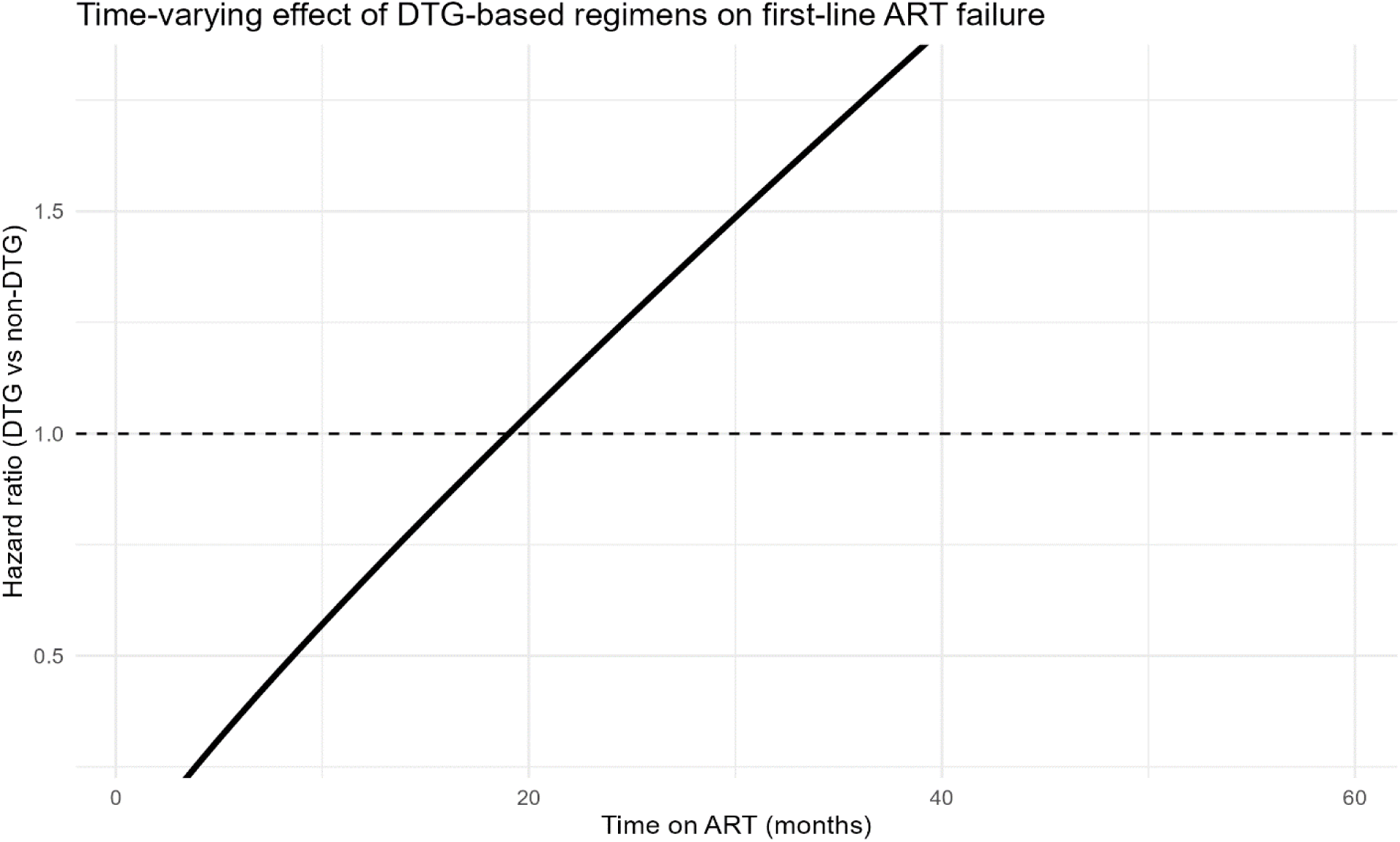
**Time-varying hazard ratio for DTG-based versus non-DTG first-line regimens.**

**Table 3:** Time-specific hazard ratios for DTG-based versus non-DTG regimens derived from the time-varying Cox model.

| Time since ART initiation (months) | Hazard ratio | 95% CI |
| --- | --- | --- |
| 6 | 0.37 | 0.22-0.60 |
| 12 | 0.67 | 0.50-0.90 |
| 24 | 1.22 | 0.88-1.70 |

**Notes:**
Hazard ratios are derived from a Cox model with a time-varying DTG effect specified as a function of $\log(\text{time})$ . Confidence intervals are calculated using the delta method.

DTG-based regimens were associated with a strong protective effect early after ART initiation (HR 0.37 at 6 months), which persisted but was attenuated at 12 months (HR 0.67). By 24 months, the hazard ratio exceeded unity (HR 1.22), with confidence intervals spanning one, indicating no apparent protective effect at later follow-up.

Higher average adherence values were observed among individuals with longer follow-up duration, reflecting the cumulative nature of the adherence measure. Because adherence was calculated across the observation period, it was not included as a baseline predictor in the primary survival models.

## 4.0 Discussion

This national cohort analysis provides longitudinal evidence on time to first-line antiretroviral therapy (ART) failure in Tanzania, using a time-to-event framework and addressing a critical gap in the predominantly cross-sectional literature on treatment outcomes in sub-Saharan Africa. By modelling time from ART initiation to virological failure, our findings align with a growing body of work demonstrating that ART durability and failure risk evolve over time and are best captured using survival analysis rather than point-prevalence measures. Recent analyses of the national HIV registry have also demonstrated the value of survival-based approaches for understanding longitudinal engagement patterns in routine programme data [16].

Several studies from Ethiopia have established the utility of time-to-failure approaches in both pediatric and adult populations. In northern Ethiopia, Shishay et al. reported a median time to treatment failure of at least 75 months among children receiving first-line ART, underscoring the potential for prolonged regimen durability when virological monitoring and adherence support are sustained [17]. Similarly, Enderis et al. employed a time-to-event design to identify predictors of first-line ART failure among adults in southern Ethiopia, demonstrating that survival analysis can reveal delayed failures that may be overlooked in cross-sectional assessments [18]. Zenebe et al. further reinforced this methodological approach by explicitly modelling time to first-line ART failure among children in Shashemene Town using Cox proportional hazards models [14].

Beyond single-country studies, the IeDEA Paediatric Cohort provided multicountry evidence on time to first-line ART failure and time to second-line switch across diverse African settings, including East Africa [19]. That study highlighted substantial heterogeneity in failure timing and emphasised the importance of longitudinal follow-up to understand treatment trajectories.

Methodologically similar approaches have also been applied in adult Ethiopian cohorts, where Agegnehu et al. defined survival time as the interval from ART initiation to virological failure and reported incidence rates and predictors using retrospective follow-up designs [20]. Similar survival-based evidence has also been reported from Addis Ababa, where Masresha Wondimgezahu and Tefera documented the incidence and predictors of treatment failure among adults receiving first-line ART [21].

Within this context, our study extends the literature by evaluating regimen-specific differences in first-line ART durability at a national scale in Tanzania. In baseline Cox models, dolutegravir (DTG)-based regimens were associated with a lower hazard of first-line failure compared with non-DTG regimens. However, the difference relative to efavirenz-based therapy did not reach conventional levels of statistical significance after adjustment. In contrast, nevirapine-based regimens consistently demonstrated a higher risk of failure, a finding that aligns with earlier Tanzanian studies reporting poorer virological outcomes and higher rates of resistance-associated mutations among patients receiving older non-nucleoside reverse transcriptase inhibitor-based regimens [5,7,10,11].

A key contribution of this analysis was the modelling of a time-varying effect for DTG, which revealed a clear temporal pattern in treatment failure risk. DTG-based regimens conferred substantial protection against virological failure during the early treatment period, with markedly lower hazards at six and twelve months after ART initiation. However, this protective effect attenuated over time, suggesting that longer-term outcomes may increasingly reflect adherence dynamics, programme transitions, or emerging resistance patterns that are now beginning to be documented in Tanzanian cohorts receiving dolutegravir-based therapy [22,23]. Pharmacy refill adherence has previously been shown to be strongly associated with virological outcomes in Tanzanian programmatic data, supporting its use as a pragmatic proxy for treatment performance in routine settings [24].

Concerns regarding delayed virological failure and resistance accumulation with longer treatment duration have been reported in both pediatric and adult cohorts in Tanzania. Studies among children and adolescents have documented the emergence of drug resistance and therapeutic failure over extended follow-up, particularly in the context of suboptimal adherence and limited regimen options [12,13]. Similarly, national and subnational data indicate that virological failure remains common among adults on both first- and second-line therapy, with incidence rates rising over time [9,15]. Our findings are therefore consistent with regional evidence suggesting that early treatment success does not guarantee long-term durability, even with high-barrier regimens such as DTG.

The observed time-varying effect also helps reconcile discrepancies between cross-sectional surveys and longitudinal cohort studies of DTG effectiveness. Cross-sectional analyses may overestimate long-term regimen performance by disproportionately capturing early treatment periods characterized by rapid viral suppression. In contrast, survival analyses incorporating extended follow-up provide a more nuanced understanding of regimen durability and failure dynamics. These results support recent calls for longitudinal monitoring of DTG outcomes, particularly as national programmes continue to expand treatment coverage and cohorts mature.

Despite the large sample size and national scope, the application of both baseline and time-varying survival models strengthens the robustness and policy relevance of our findings. These findings should be interpreted in light of the limitations of routinely collected programmatic data, including incomplete clinical covariates and potential misclassification of virological failure.

In conclusion, this study reinforces the importance of time-to-event approaches for evaluating first-line ART outcomes. It provides new evidence on the temporal dynamics of DTG effectiveness in a national programme setting. While DTG-based regimens offer strong early protection against virological failure, sustained long-term success will depend on continued adherence support, resistance surveillance, and differentiated care strategies as treatment duration increases. These findings have direct implications for ART programme monitoring in Tanzania and similar settings, underscoring the need to move beyond cross-sectional metrics when assessing regimen performance.

## Limitations

This study has several limitations that should be considered when interpreting the findings. First, the analysis relied on routinely collected national programmatic data, which were not initially designed for research purposes. As a result, some clinically important variables, including baseline CD4 count, WHO clinical stage, tuberculosis co-infection, and baseline viral load, were incompletely recorded and could not be incorporated into multivariable models. Residual confounding by unmeasured clinical factors, therefore, remains possible.

Second, virological failure was defined as the first documented viral load ≥1,000 copies/mL occurring at least 6 months after ART initiation. Although this definition aligns with national and WHO guidelines, confirmatory viral load measurements following enhanced adherence counselling were not consistently available in the dataset. Consequently, some transient viraemia may have been misclassified as treatment failure.

Third, adherence was measured using pharmacy refill data, which may not fully capture actual medication intake and may overestimate true adherence. Fourth, patients who died, transferred out, or were lost to follow-up before six months of ART initiation were excluded, which may have resulted in underestimation of early treatment failure.

Finally, due to computational constraints, analyses were conducted on a large simple random sample drawn from the national CTC-2 database rather than the full dataset. However, the sampling approach ensured equal probability of selection prior to application of eligibility criteria, supporting the internal validity of the findings.

## 5.0 Conclusions and Recommendations

In this large national cohort of people living with HIV in Tanzania, the risk of first-line antiretroviral therapy failure varied significantly by regimen type and patient characteristics. Dolutegravir-based regimens were associated with a lower risk of treatment failure compared with non-DTG regimens during early follow-up, although this protective effect attenuated over time. Nevirapine-based regimens were consistently associated with a higher risk of failure relative to efavirenz-based therapy. Younger age and female gender were associated with higher failure hazard in the baseline survival models.

These findings reinforce the continued programmatic prioritization of DTG-based first-line regimens while highlighting the need for sustained virological monitoring and continued support for treatment continuity beyond the initial months of therapy. Enhanced viral load monitoring, particularly during longer-term follow-up, is critical for detecting emerging treatment failure among patients on DTG-based therapy. Targeted interventions addressing retention in care and virological monitoring, especially among younger individuals and women, may help sustain long-term treatment success.

Future studies should integrate more complete clinical and laboratory data, including baseline immunological status and co-morbidities, to further refine risk stratification and inform individualized HIV care strategies.

## Declarations

### Ethics approval and consent to participate

Ethical approval for this study was obtained from the Directorate of Research, Publications and Innovation at Muhimbili University of Health and Allied Sciences (MUHAS) (Reference: DA.25/111/28/01/2021). Permission to access and analyze programmatic HIV care and treatment data was granted by the National AIDS, Sexually Transmitted Infections and Hepatitis Control Programme (NASHCoP). The study used routinely collected, fully anonymized secondary data; therefore, individual informed consent was waived in accordance with national ethical guidelines and institutional review board regulations.

## Consent for publication

Not applicable.

## Availability of data and materials

The data used in this study are derived from the national Care and Treatment Clinic (CTC-2) database managed by NASHCoP. Due to legal and ethical restrictions related to patient confidentiality and data governance, the datasets are not publicly available. De-identified data may be made available from NASHCoP upon reasonable request and subject to relevant institutional approvals.

## Authors’ contributions

JM, HB, and RZS conceived and designed the study. JM and RZS conducted data cleaning and statistical analyses. RZS provided methodological oversight and supervised the analysis. VS and PN facilitated access to the CTC-2 data and provided programmatic context. JM and RZS drafted the manuscript. All authors critically reviewed the manuscript, contributed to its intellectual content, and approved the final version for publication.

## Acknowledgements

The authors acknowledge the National AIDS, Sexually Transmitted Infections and Hepatitis Control Programme (NASHCoP) for granting access to the CTC-2 database. We also acknowledge the dedication of healthcare workers at HIV CTCs across Tanzania to data collection and patient care.

## References

1. World Health Organization. HIV data and statistics. 2025 [cited 11 Mar 2026]. Available: https://www.who.int/teams/global-hiv-hepatitis-and-stis-programmes/hiv/strategic-information/hiv-data-and-statistics

2. United Republic of Tanzania. Tanzania HIV Impact Survey 2022-2023. 2024 [cited 18 Dec 2025]. Available: https://www.nbs.go.tz/nbs/takwimu/THIS2022-2023/THIS2022-2023_Summary_Sheet.pdf

3. The United Republic of Tanzania. National Guidelines for Management of HIV and AIDS. 2019 [cited 11 Mar 2026]. Available: https://hivpreventioncoalition.unaids.org/en/resources/tanzania-national-guidelines-management-hiv-and-aids-7th-edition

4. Ministry of Health. National Guidelines for the management of HIV and AIDS in Tanzania. 2017 [cited 11 Mar 2026]. Available: https://platform.who.int/docs/default-source/mca-documents/policy-documents/guideline/TZA-RH-43-02-GUIDELINE-2017-eng-National-Guidelines-for-Management-of-HIV-and-AIDS-2017-AR.pdf

5. Hawkins C, Ulenga N, Liu E, Aboud S, Mugusi F, Chalamilla G, et al. HIV virological failure and drug resistance in a cohort of Tanzanian HIV-infected adults. Journal of Antimicrobial Chemotherapy. 2016;71: 1966–1974. doi:10.1093/jac/dkw051

6. Jaka HM, Mshana SE, Liwa AC, Peck R, Kalluvya S. Prevalence of immunological failure and durability of first line antiretroviral therapy at Bugando Hospital Mwanza, Tanzania. Tanzania Medical Journal. 2010;24. doi:10.4314/tmj.v24i2.53278

7. Henerico S, Mikasi SG, Kalluvya SE, Brauner JM, Abdul S, Lyimo E, et al. Prevalence and patterns of HIV drug resistance in patients with suspected virological failure in North-Western Tanzania. Journal of Antimicrobial Chemotherapy. 2022;77: 483–491. doi:10.1093/jac/dkab406

8. Sangeda RZ, Gómes P, Rhee S-Y, Mosha F, Camacho RJ, Van Wijngaerden E, et al. Development of HIV Drug Resistance in a Cohort of Adults on First-Line Antiretroviral Therapy in Tanzania during the Stavudine Era. Microbiol Res (Pavia). 2021;12: 847–861. doi:10.3390/microbiolres12040062

9. Mwavika ET, Kunambi PP, Masasi SJ, Lema N, Kamori D, Matee M. Prevalence, rate, and predictors of virologic failure among adult HIV-Infected clients on second-line antiretroviral therapy (ART) in Tanzania (2018-2020): a retrospective cohort study. Bull Natl Res Cent. 2024;48: 96. doi:10.1186/s42269-024-01248-5

10. Ngarina M, Kilewo C, Karlsson K, Aboud S, Karlsson A, Marrone G, et al. Virologic and immunologic failure, drug resistance and mortality during the first 24 months postpartum among HIV-infected women initiated on antiretroviral therapy for life in the Mitra plus Study, Dar es Salaam, Tanzania. BMC Infect Dis. 2015;15: 175. doi:10.1186/s12879-015-0914-z

11. Samizi FG, Panga OD, Mulugu SS, Gitige CG, Mmbaga EJ. Rate and predictors of HIV virological failure among adults on first-line antiretroviral treatment in Dar Es Salaam, Tanzania. The Journal of Infection in Developing Countries. 2021;15: 853–860. doi:10.3855/jidc.13603

12. Muri L, Gamell A, Ntamatungiro AJ, Glass TR, Luwanda LB, Battegay M, et al. Development of HIV drug resistance and therapeutic failure in children and adolescents in rural Tanzania. AIDS. 2017;31: 61–70. doi:10.1097/QAD.0000000000001273

13. Maseke I, Joachim A, Kamori D, Abade A, Moremi N, Majigo M. Prevalence of human immunodeficiency virus drug resistance and factors associated with high viral load among adolescents on antiretroviral therapy in Dar Es Salaam, Tanzania. HIV Res Clin Pract. 2024;25. doi:10.1080/25787489.2024.2400827

14. Zenebe E, Washo A, Addis Gesese A. Time to First-Line Antiretroviral Treatment Failure and Its Predictors among HIV-Positive Children in Shashemene Town Health Facilities, Oromia Region, Ethiopia, 2019. Karwowski J, editor. The Scientific World Journal. 2021;2021: 1–9. doi:10.1155/2021/8868479

15. Rugemalila J, Kamori D, Maokola W, Mizinduko M, Barabona G, Masoud S, et al. Acquired HIV drug resistance among children and adults receiving antiretroviral therapy in Tanzania: a national representative survey protocol. BMJ Open. 2021;11: e054021. doi:10.1136/bmjopen-2021-054021

16. Mwakyomo J, Sangeda RZ, Mushi H, Njau P. Time to registry discontinuity in Tanzania’s national HIV care registry: a survival analysis of population mobility patterns. 2026. pp. 1–23. doi:10.64898/2026.03.07.26347830

17. Shishay AS, Gebrezgiabher BB, Gebreyohannes YK, Negash BM, Tewele BG, Berhe ES, et al. Time to treatment failure and its predictors among children receiving first-line antiretroviral therapy in Tigray Region public general hospitals, North Ethiopia, 2024: Retrospective cohort study. Evans D, editor. PLoS One. 2026;21: e0339269. doi:10.1371/journal.pone.0339269

18. Oumer Enderis B, Hussen Hebo S, Kote Debir M, Boti Sidamo N, Shegaze Shimber M. Predictors of Time to First Line Antiretroviral Treatment Failure among Adult Patients Living with HIV in Public Health Facilities of Arba Minch Town, Southern Ethiopia. Ethiop J Health Sci. 2019;29. doi:10.4314/ejhs.v29i2.4

19. Wools-Kaloustian K, Marete I, Ayaya S, Sohn AH, Van Nguyen L, Li S, et al. Time to First-Line ART Failure and Time to Second-Line ART Switch in the IeDEA Pediatric Cohort. JAIDS Journal of Acquired Immune Deficiency Syndromes. 2018;78: 221–230. doi:10.1097/QAI.0000000000001667

20. Agegnehu CD, Merid MW, Yenit MK. Incidence and predictors of virological failure among adult HIV patients on first-line antiretroviral therapy in Amhara regional referral hospitals; Ethiopia: a retrospective follow-up study. BMC Infect Dis. 2020;20: 460. doi:10.1186/s12879-020-05177-2

21. Masresha Wondimgezahu S, Tefera T. Incidence and Predictors of Treatment Failure among Adult on First-Line Antiretroviral Therapy in Gullele Sub City, Addis Ababa, Ethiopia 2023: A Retrospective Study in Selected Public Health Facilities. International Journal of Virology and AIDS. 2023;10. doi:10.23937/2469-567X/1510092

22. Kilapilo MS, Sangeda RZ, Bwire GM, Sambayi GL, Mosha IH, Killewo J. Adherence to Antiretroviral Therapy and Associated Factors Among People Living With HIV Following the Introduction of Dolutegravir Based Regimens in Dar es Salaam, Tanzania. Journal of the International Association of Providers of AIDS Care (JIAPAC). 2022;21: 232595822210845. doi:10.1177/23259582221084543

23. Bwire GM, Aiko BG, Mosha IH, Kilapilo MS, Mangara A, Kazonda P, et al. High viral suppression and detection of dolutegravir-resistance associated mutations in treatment-experienced Tanzanian adults living with HIV-1 in Dar es Salaam. Sci Rep. 2023;13: 20493. doi:10.1038/s41598-023-47795-1

24. Lugoba MD, Sangeda RZ, Mutagonda RF, Mwakyomo J, Musiba G, Sambu V, et al. Leveraging Machine Learning Models and Pharmacy Refill Adherence as a Cost-Effective Proxy for Predicting HIV Viral Suppression during Antiretroviral Therapy in Resource-Limited Settings. 2026. doi:10.64898/2026.01.05.26343496

